# Cardiovascular Risk in BRCA1/2 Mutation Carriers: A Matched Cohort Study of Breast Cancer Survivors

**DOI:** 10.64898/2026.07.26.26350298

**Authors:** Mehdi Dehghan Manshadi, Narges Manouchehri, Laila Hubbert, Annelie Liljegren, Ali Manouchehrinia, Christina Linder-Stragliotto, Johanna Rantala, Elham Hedayati, Narsis A.Kiani

## Abstract

**Introduction:** Cardiovascular disease (CVD) is a leading non-cancer cause of morbidity among breast cancer (BC) survivors. Among them, women carrying germline BRCA1 or BRCA2 mutations (BRCA-BC) may be at particular risk of CVD, but evidence is inconsistent. The objective of this study is to determine whether BRCA-BC independently influences the risk for CVD after BC diagnosis in the Stockholm-Gotland region in Sweden (2008-2019).

**Methods:** In this registry-based cohort study, we used exact matching on age at diagnosis, tumor stage, laterality, and pre-existing CVD or risk factors to construct 32 matched (1:1) subgroups. Multi-state Cox proportional hazards models estimated hazard ratios (HRs) for transitions from BC diagnosis to first cardiovascular event, while accounting for competing risks of distant metastasis or non-cardiovascular death.

**Results:** In matched subgroups, BRCA-BC experienced fewer CVD (6.4% vs. 11.2% (IQR 9.4%-12.2%), but significantly more competing events (22.3% vs. 10.1% (IQR 8.9%-11.3%); *p*<0.05. Multi-state Cox models revealed an inverse association between BRCA-BC status and the first cardiovascular event (HR<1), but a higher hazard of the competing risk. Cardiovascular events clustered in the first year after BC diagnosis, especially among BRCA-BC, suggesting truncated time at risk.

**Conclusion:** BRCA-BC did not demonstrate increased cardiovascular risk after BC diagnosis. The apparent inverse association with CVD likely reflects the high incidence of competing risks, which limit the window for CVD to manifest. A small subgroup of long-term BRCA-BC survivors may represent biologically distinct individuals with different cardiovascular susceptibility, warranting further investigation.

## Introduction

Cardiovascular disease (CVD) has emerged as a major competing cause of morbidity and mortality among breast cancer (BC) survivors. Improvements in early detection and multimodal therapy have substantially increased survival across stages I-III, shifting the clinical focus from short-term cancer outcomes toward the long-term sequelae of treatment. Anthracycline- and trastuzumab-based regimens, chest radiotherapy, and certain endocrine therapies are well recognized for their cardiovascular toxicity potential, and accumulating data suggest that BC survivors face elevated risks of heart failure, ischemic heart disease, and stroke compared to the general population [1,2]. In addition, BC and CVD share major, often modifiable risk factors such as age, obesity, hypertension, dyslipidemia, diabetes, and tobacco use, as well as common pathobiological pathways (e.g., chronic inflammation and metabolic dysregulation), which may amplify therapy-related cardiotoxicity. Emerging work highlights inflammatory mediators such as S100A8/9 and the NLRP3 inflammasome as shared mechanisms linking tumor biology and vascular injury, further narrowing the boundary between cancer and CVD [3,4]. Moreover, the period immediately following BC diagnosis and treatment initiation is characterized by intense physiological stress and rapid treatment exposure, during which clustering of cardiovascular events (CVD and cardiovascular death) has been observed in several cohorts [5–7]. These observations underscore the importance of identifying patient subgroups that may be particularly vulnerable to cardiovascular complications [8].

Women with germline BRCA1 or BRCA2 mutations who are diagnosed with BC (BRCA-BC) represent such a cohort of interest. BRCA mutations account for approximately 5-10% of all BC cases and confer high lifetime risks of breast and ovarian cancer. Clinically, these patients are characterized by younger age at diagnosis, higher prevalence of triple-negative tumors, and a tendency toward more aggressive disease biology [9–11]. As a result, BRCA-BC frequently undergo intensive multimodal treatments, including alkylating chemotherapy, platinum agents, and bilateral mastectomy or oophorectomy at earlier ages than their sporadic counterparts. These features alone raise the possibility of heightened risk of cardiovascular toxicity. Beyond oncologic features, mechanistic evidence suggests that BRCA1 and BRCA2 play roles in cardiovascular functions in addition to their tumor-suppressive functions.

Preclinical studies have demonstrated that cardiomyocyte-specific BRCA1 deletion compromises DNA damage repair, impairs mitochondrial integrity, and exacerbates anthracycline-induced cardiomyopathy [12–16]. Similarly, endothelial cell-specific loss of BRCA2 has been linked to accelerated atherogenesis and dysregulation of oxidative stress pathways [17]. These mechanistic data align with recent reviews emphasizing DNA repair and mitochondrial signaling as central mechanisms in cancer therapy-related cardiovascular toxicity [14]. Together, they provide a biological rationale for hypothesizing that BRCA1/2 mutation may sensitize the cardiovascular system to therapy-related and endogenous stressors.

Some reports suggest an association between BRCA mutations and increased cardiovascular morbidity or mortality, potentially mediated by surgical menopause, risk-reducing surgery, or repeated exposure to DNA-damaging agents [18, 19]. Others, however, including prospective assessments of cardiovascular fitness and function in BRCA-BC, have found no meaningful differences compared to non-mutation carriers [20,13]. Several methodological limitations likely contribute to these discrepancies. Many studies focused exclusively on clinically tested BRCA1/2 mutations, while comparison cohorts often included patients with unknown genetic status, raising the possibility of contamination by undiagnosed mutation carriers. Few investigations captured cardiovascular outcomes in the early period after BC diagnosis (the first 12 months), when risk is highest, and most did not adequately account for competing risks such as distant metastasis or non-cardiovascular death. Failure to address these competing events may lead to underestimation of risk for CVD in BRCA-BC, who frequently experience earlier recurrence and mortality, thereby shortening the period during which cardiovascular events can manifest. Moreover, rigorous adjustment for confounding variables, including pre-existing CVD, established cardiovascular risk factors (CVRFs), such as hypertension and diabetes, and tumor stage, has seldom been applied in this setting. Recent reviews in cardio-oncology further emphasize the importance of integrating genetic predisposition and health metrics (e.g. blood pressure, cholesterol levels and long-term blood sugar) into risk for CVD prediction among cancer survivors [21,22].

Against this background, robust comparative data are needed to clarify whether BRCA-BC differ from sporadic BC in their cardiovascular risk following BC diagnosis. To address this gap, we conducted a registry-based matched cohort study within the Stockholm-Gotland region in Sweden, leveraging detailed clinical, treatment, and outcome data. By constructing multiple matched subgroups of BRCA-BC and sporadic BC, and applying multi-state Cox models that explicitly account for competing events, we aimed to provide an estimate of the short- to intermediate-term risk for CVD associated with BRCA-BC status. This design enables evaluation of whether BRCA-BC status independently influences cardiovascular outcomes among breast cancer survivors, thereby contributing evidence to an area of clinical and biological uncertainty.

## Methods

### Study cohort

In this register-based cohort analysis, we identified 17,443 women diagnosed with BC (stages I-III and ductal carcinoma in situ [DCIS]) in the Stockholm-Gotland region of Sweden between January 1, 2008, and December 31, 2019. Among these, 233 individuals (1.3%) with a confirmed pathogenic BRCA1/2 variant were classified as BRCA-BC. The remaining 17,210 women (98.7%) had no record of a pathogenic BRCA1/2 variant in regional or national registries and were therefore considered the reference group (hereafter referred to as Sporadic-BC). Data from regional and national quality registries provided detailed information on pre-existing CVD, CVRFs, cancer treatments, and demographic characteristics. The data sources and linkage procedures have been described previously [23]. Cardiovascular outcomes were identified from both inpatient and outpatient records using International Classification of Diseases (ICD) codes and registry-defined interventions. For each patient, the outcome of interest was defined as the earliest CVD or cardiovascular death occurring after the date of BC diagnosis and before the occurrence of distant metastasis. The outcomes in the “early period” after breast cancer diagnosis refer to those occurring within the first 12 months following the initial diagnosis.

To minimize confounding, each BRCA-BC patient was matched (1:1) with Sporadic-BC on age at diagnosis, tumor stage, tumor laterality, and the presence of pre-existing CVD and CVRFs. Only individuals with at least one eligible match were retained, resulting in a small, balanced analytic subgroup (Figure 1).

**Figure 1:**
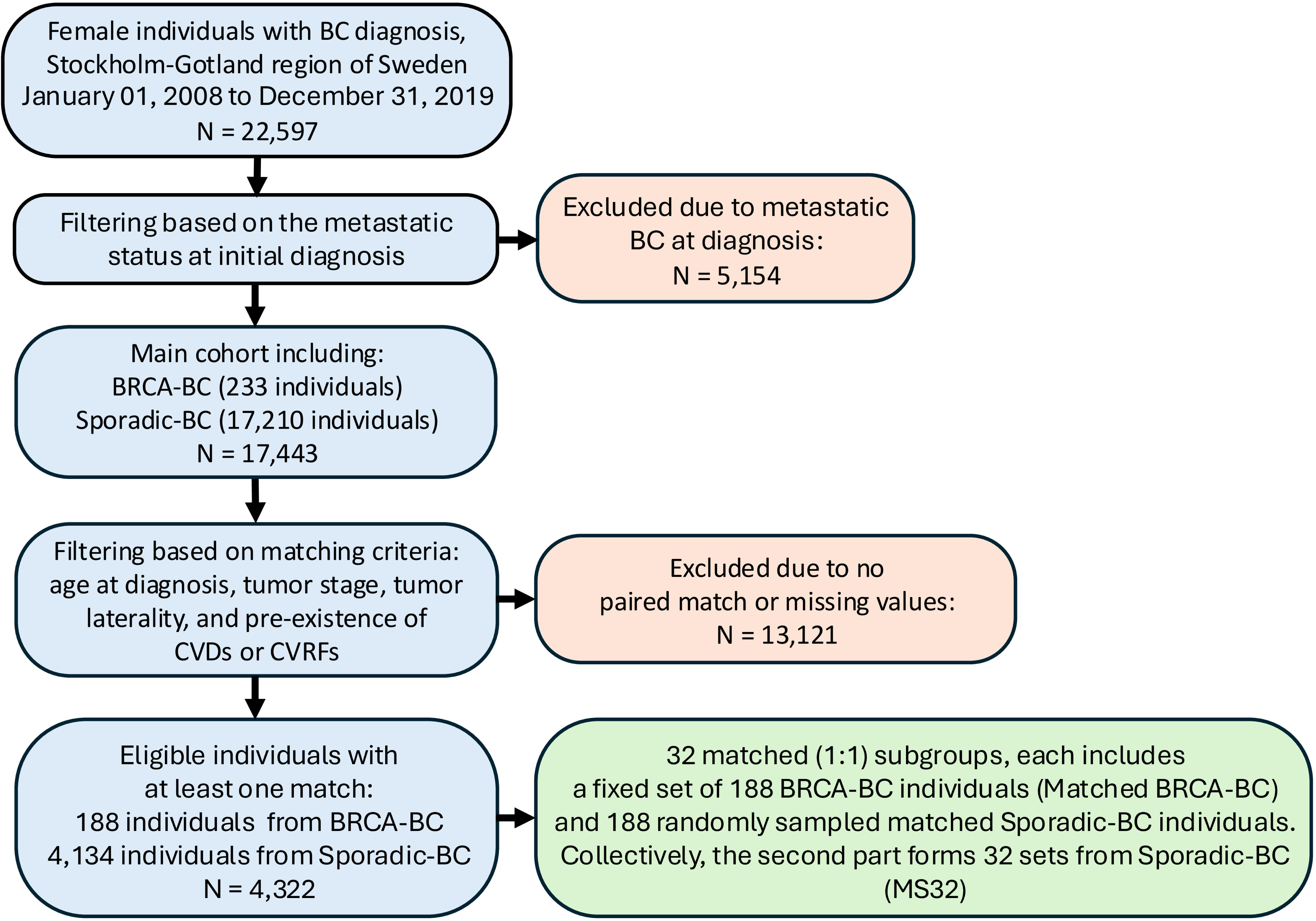
Framework for the matching process. In this study, individuals with known positive metastatic status at initial diagnosis were excluded. Of the 17,443 remaining individuals, the matching process identified 4,322 eligible individuals, from which we generated matched subgroups (1:1). BC=Breast cancer, BRCA-BC=Individuals with BRCA1/2 germline mutations who diagnosed with breast cancer in the main population, N=number of individuals, Sporadic-BC=Individuals with breast cancer without known status of BRCA1/2 mutation.

This study was approved by the Regional Ethics Review Board (2018/2669-31/2) and adhered to the STROBE guidelines [24] and the Declaration of Helsinki [25]. According to Swedish legislation, written informed consent is not required from individuals included in national quality registries for the use of their data in healthcare research. The authors acknowledge assistance from an AI-based language tool (ChatGPT, OpenAI) used solely for language refinement.

### Cardiovascular risk factors and morbidity

CVRFs and CVD were identified using ICD-10 codes. CVRFs included diagnoses of hypertension (I10) and diabetes mellitus (E10, E11). Although hypertension is classified as a cardiovascular disease, chronically elevated blood pressure causes endothelial injury, arterial stiffness/atherosclerosis, and cardiac remodeling, thereby raising the risk of MI, stroke, heart failure, and arrhythmias. Accordingly, in our study, we treat hypertension as a risk factor rather than an outcome.

CVD was defined as one or more of the following: Ischemic heart diseases (I20-25) and Myocardial infarction (I21), arrhythmias including atrial fibrillation/flutter (AF) (I48), Atrioventricular block (AV-block III) (I44.2), ventricular tachycardia (VT) (I47.2), ventricular fibrillation (VF), (I49.0), cardiac arrest (I46), conduction disorders, paroxysmal tachycardia, other cardiac arrhythmias (I44, I45, I47, I49), heart failure (HF) (I50), cardiomyopathy (I42, I43), valvular diseases (I34-I37), and acute/primary cerebrovascular diseases (I60-I67), including cerebrovascular accident (CVA) (I63, I64), additional vascular disorders as atherosclerosis (I70), pulmonary embolism (I26), venous thrombosis/embolism (I81-I82) was also reported.

Cardiovascular interventions, including coronary stent implantation (Z95.5), coronary artery bypass grafting (CABG) (Z95.1), pacemaker implantation (Z95.0), and cardiac resynchronization therapy or implantable cardioverter defibrillator (CRT/ICD) (Z95.810), were also included in the definition of CVD.

### Handling of missing information

To address missing data on chemotherapy treatment, we used the Swedish Prescribed Drug Register [26]. Prescription of supportive medications commonly used in chemotherapy care was used as a proxy for treatment. We considered prescriptions from diagnosis to four months post-surgery (if performed) as indicative of chemotherapy. Details of selected medications and their Anatomical Therapeutic Chemical classification system (ATC codes) are provided in the Supplemental Table 1.

By applying the previously described strategy using the Swedish Prescribed Drug Register, we were able to infer chemotherapy treatment for 3,219 individuals (18.5%) with missing treatment data. To validate this approach, we compared the inferred data with available records of actual chemotherapy administration, demonstrating high concordance. For cases with missing radiotherapy administration data, planned radiotherapy was used as a proxy for exposure. Furthermore, individuals with no documented CVD during follow-up were assumed not to have experienced any CVD.

### Statistical analysis

Baseline clinical and demographic characteristics were compared between the cohorts using Student’s t-test for continuous variables and the chi-square (χ²) test for categorical variables.

Hazard ratios (HR) were estimated using multi-state Cox proportional hazards models. Models were adjusted for the following covariates: receipt of chemotherapy (yes/no), receipt of radiotherapy (yes/no), tumor laterality (right vs. left/bilateral), and tumor stage (I/DCIS vs. II/III). Relapse status (yes/no) was modeled as a time-varying covariate, and the matching index was included as a random effect (frailty term). Pre-existing CVRFs (hypertension, diabetes) and pre-existing CVD were handled through stratification to avoid violation of the proportional hazard’s assumption. The assumption of proportionality was formally evaluated using Schoenfeld residuals.

The primary transition of interest was from BC diagnosis to the first CVD or cardiovascular death. Competing events were defined as transitions from BC diagnosis to either distant metastasis or non-cardiovascular death. Patients without events were censored at the end of follow-up (December 31, 2019). The median follow-up, calculated using the reverse Kaplan-Meier method.

To address the fact that many BRCA-BC individuals had multiple eligible comparators among Sporadic-BC individuals, we constructed 32 matched (1:1) subgroups by repeatedly and randomly sampling from the pool of eligible Sporadic-BC individuals. This repeated sampling ensured that the results were not dependent on any single matching choice. By doing this, we obtained 32 matched (1:1) subgroups (376 individuals each), all containing the same 188 BRCA-BC individuals (hereafter referred to as Matched BRCA-BC) and, in each subgroup, a different randomly sampled set of 188 matched Sporadic-BC individuals; we refer to these 32 sets of matched Sporadic-BC as *MS32*.

A post hoc power calculation was performed for the primary matched subgroups. Using the observed event rate in the overall population as the basis for expected incidence, the analysis indicated approximately ∼99% power to detect a hazard ratio of 0.22 at a two-sided α of 0.05. While this approach provides a reasonable approximation, it may underestimate or overestimate true power, as matching alters both the number of events and the variance structure relative to the full cohort. Multi-state Cox proportional hazards models have been applied to the 32 matched subgroups, explicitly incorporating competing risks of distant metastasis and non-cardiovascular death.

All analyses, including preprocessing, model fitting, and visualization, were conducted in R version 4.5.1[27] and RStudio 5.1 [28]. Multi-state survival analyses were implemented using the *mstate* package [29].

## Results

Baseline demographic and Clinical data are provided in Table 1. The median age at BC diagnosis was 20.0 years younger in the BRCA-BC than in the Sporadic-BC (*p*<0.001). The median follow-up was 0.7 years longer in BRCA-BC than in Sporadic-BC, and this difference was not statistically significant (*p*=0.45). Treatment patterns also differed. Radiotherapy was less common in BRCA-BC (by 13.8%; *p*<0.001), though missing data were more frequent in BRCA-BC. The use of chemotherapy was higher in BRCA-BC (by 35.9%; *p*<0.001). At baseline, pre-existing CVD was less common among BRCA-BC (by 6.6%; *p*=0.001), and CVRFs were also less frequent (by 13.1%; *p*<0.001).

**Table 1.**
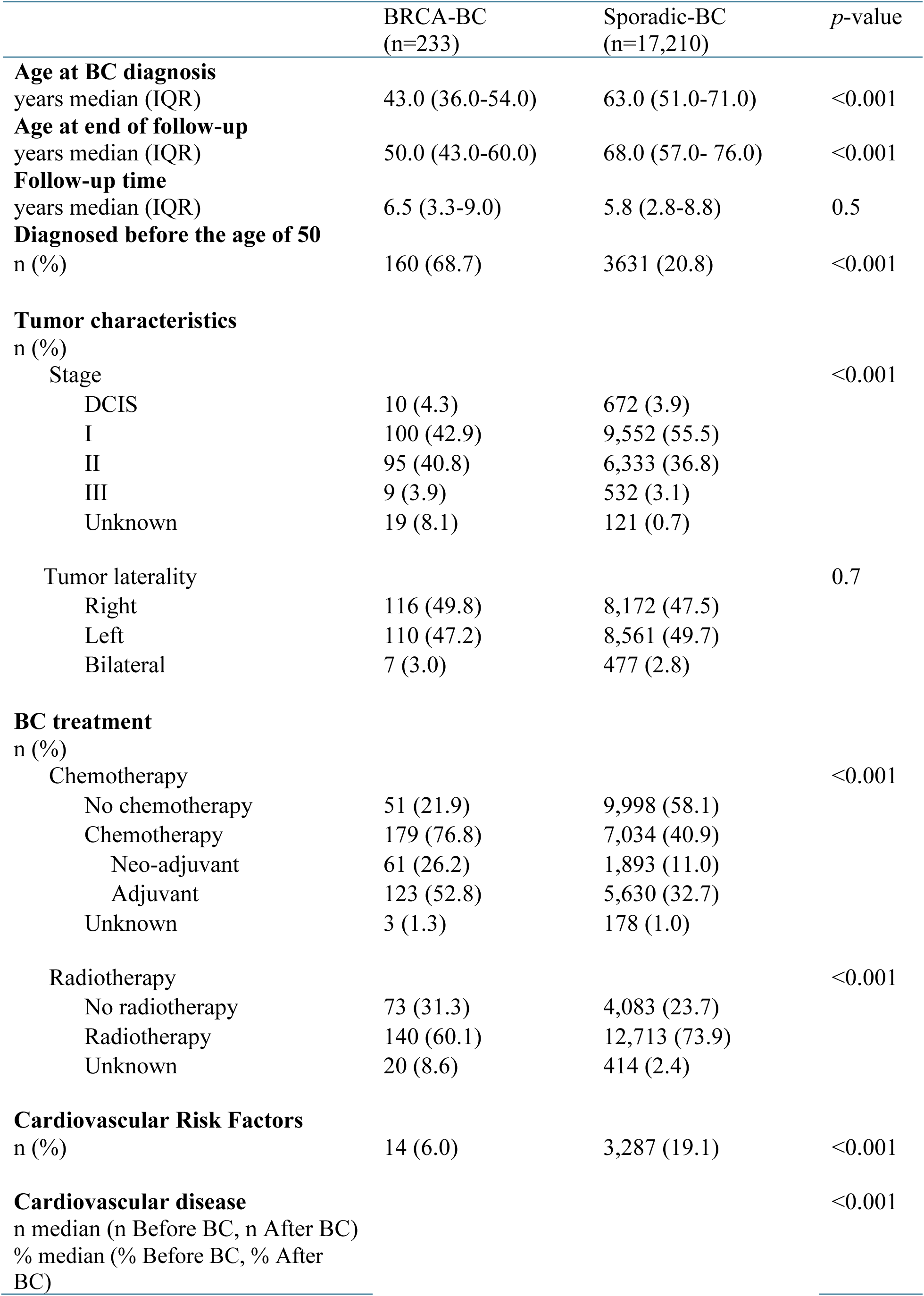
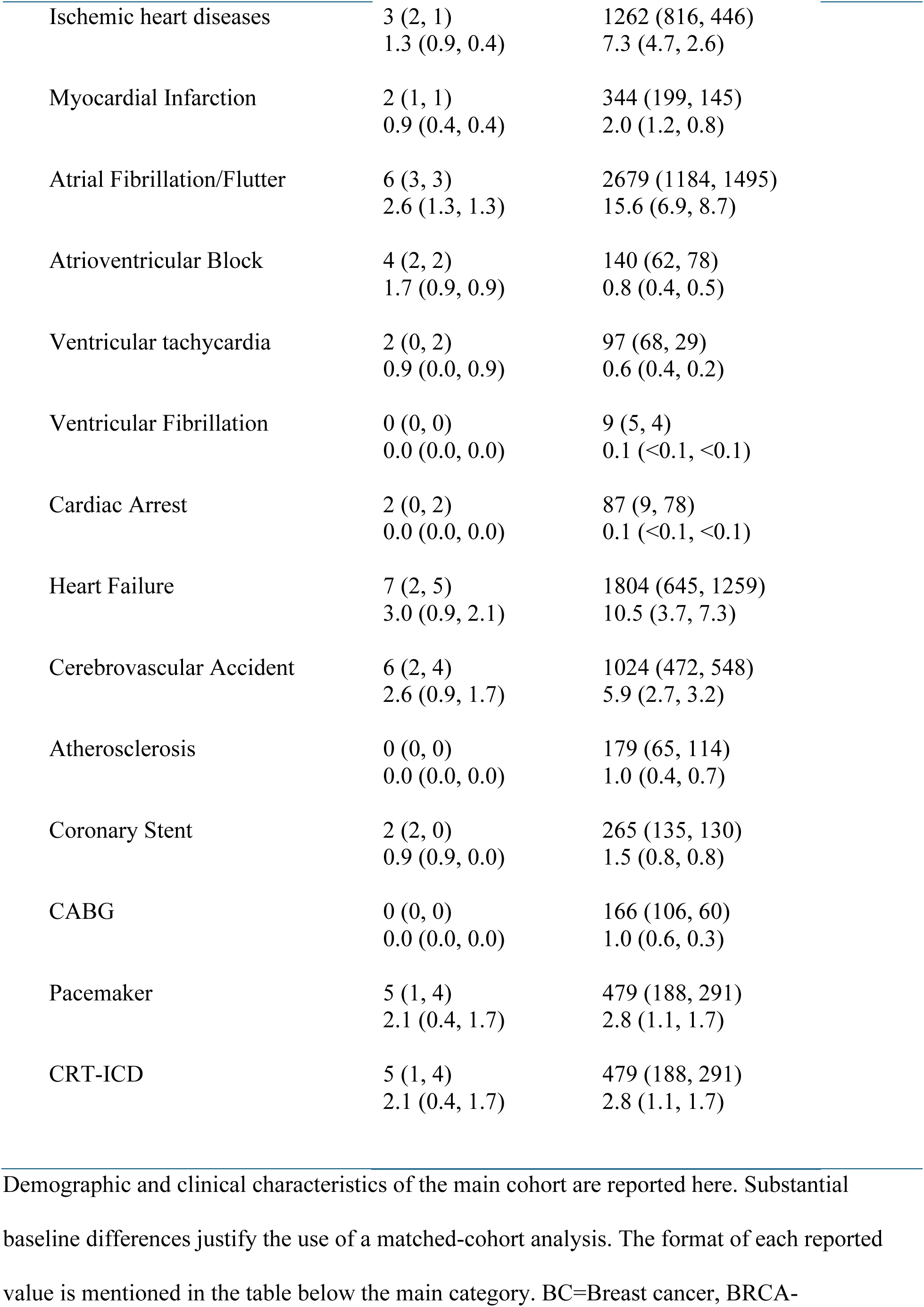
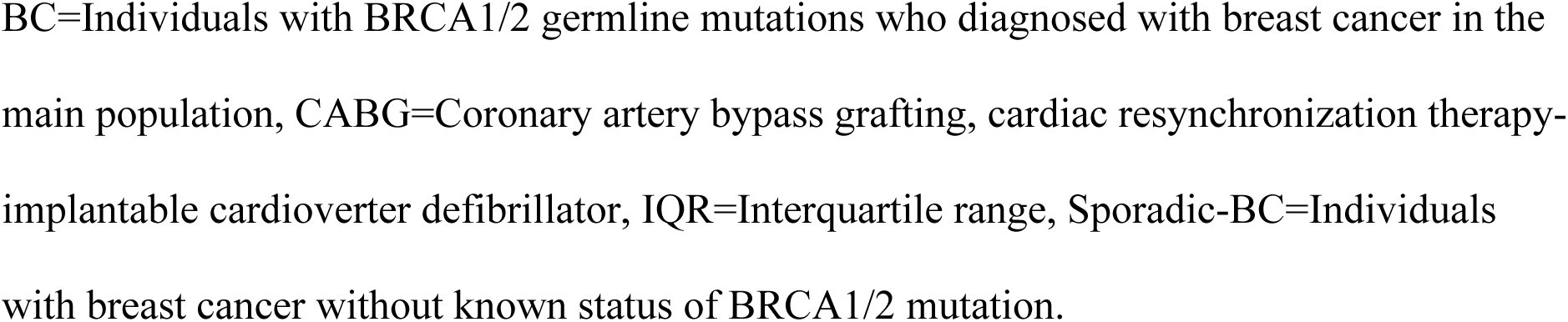
Demographic and Clinical Characteristics of the main cohort.

### Matched subgroups reveal distinct patterns of CVD and cancer progression

Exact matching on age at diagnosis, tumor stage, tumor laterality, and pre-existing CVD/CVRFs yielded a set of 4,322 eligible individuals (188 from BRCA-BC and 4,134 from Sporadic-BC). In the matched subgroups, the median follow-up was 1.3 years longer in Matched BRCA-BC than in MS32 (*p>*0.05 for all 32 subgroups), indicating no statistically significant difference in surveillance time between subgroups.

Matched BRCA-BC was more likely to receive chemotherapy (by 17.0%; *p*<0.05) and less likely to receive radiotherapy (by 13.8%; *p*<0.05). Relapse and distant metastasis occurred more frequently among Matched BRCA-BC (by 18.1% for relapse, *p<*0.05; and by 10.4% for metastasis; *p*<0.05). Despite the higher burden of aggressive disease in Matched BRCA-BC, they still showed fewer cardiovascular events, whereas competing risks of distant metastasis or non-cardiovascular death were substantially higher (4.8% fewer main risk and 12.2% higher competing risk; *p*<0.05). Across subgroups, all tests for chemotherapy, radiotherapy, and relapse were significant (*p*<0.05), whereas 29 of 32 were significant for distant metastasis and the composite competing outcome (Table 2).

**Table 2.**
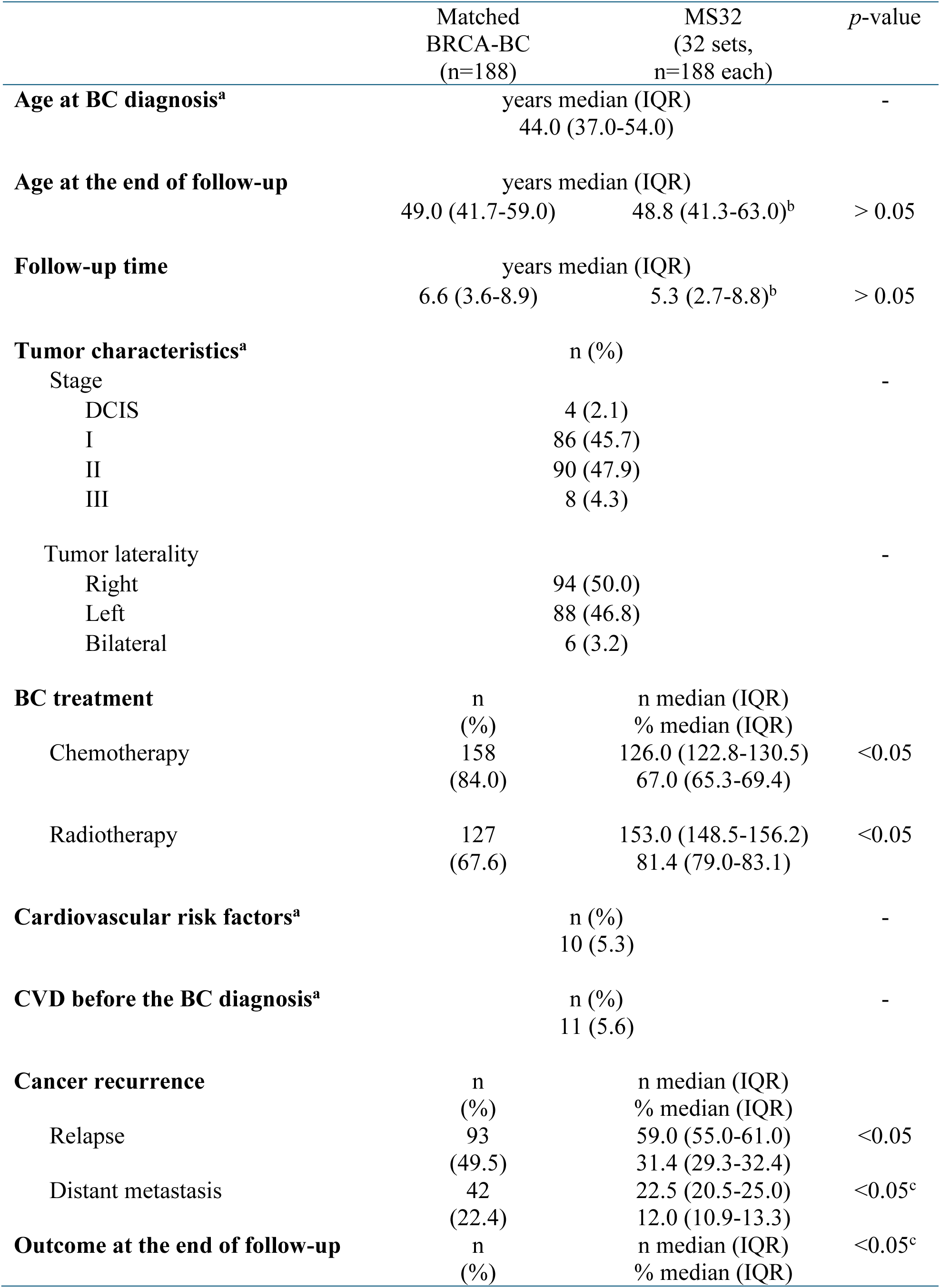
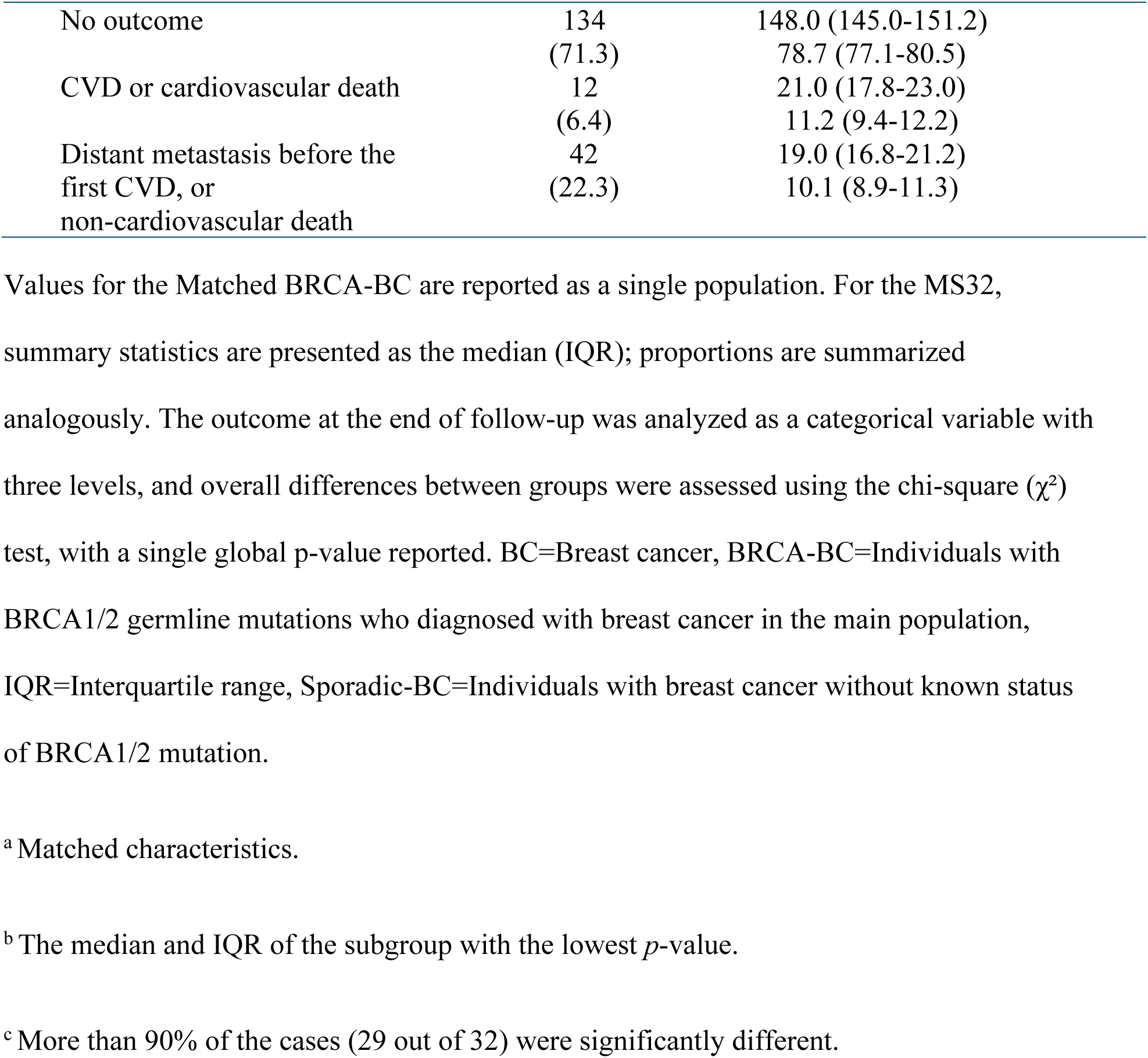
Characteristics of the matched subgroups.

### The incidence of CVD increases during the first-year post-diagnosis

In both the BRCA-BC and Sporadic-BC cohorts, the number of CVDs showed a temporal concentration during the early post-diagnosis period. Among individuals who developed CVD, the majority of first events occurred within the first 12 months after BC diagnosis (Figure 2A). This pattern was consistent across matched subgroups, demonstrating that the first year after BC diagnosis represents a period of heightened risk of cardiovascular toxicity. When restricting outcomes to CVD occurring prior to distant metastasis, the distribution of CVD or cardiovascular death in the BRCA-BC became sparse. Nearly all cardiovascular events in BRCA-BC were confined to the first year after diagnosis, with very few CVD (n=3) observed in later follow-up (Figure 2B). This suggests that the window for cardiovascular events is compressed by the high incidence of competing risks, namely metastasis or non-cardiovascular death, at younger ages in BRCA-BC.

**Figure 2:**
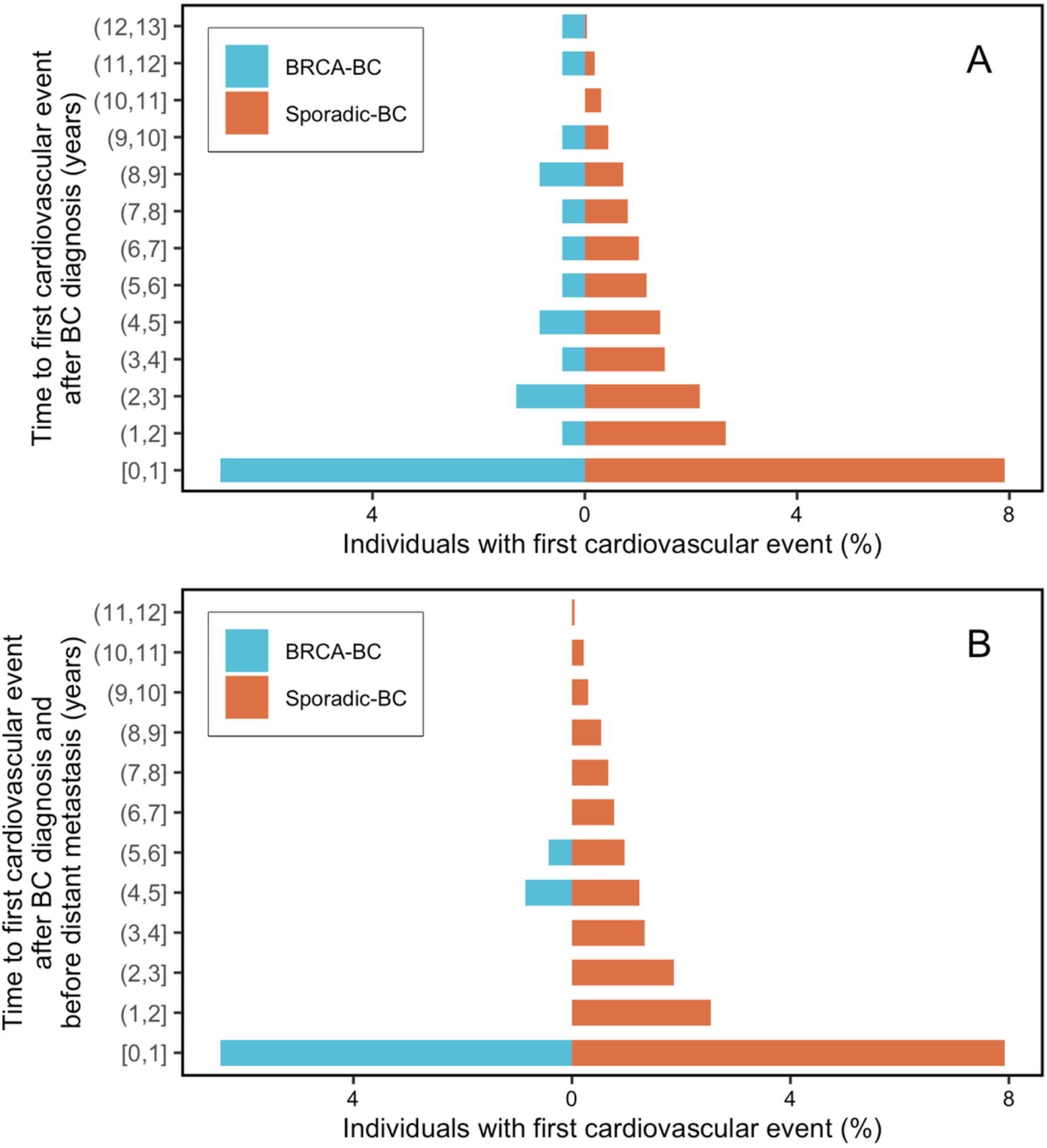
Age distribution of the first cardiovascular event after BC diagnosis. The age distribution of the first cardiovascular event after BC diagnosis is significantly different between BRCA-BC and Sporadic-BC. Both cohorts showed a temporal concentration during the early post-diagnosis period. (A) shows the age distribution of time to the first cardiovascular event, both before and after developing distant metastasis, and (B) shows only the events that occurred before developing distant metastasis. BC=Breast cancer, BRCA-BC=Individuals with BRCA1/2 germline mutations who diagnosed with breast cancer in the main population, Sporadic-BC=Individuals with breast cancer without known status of BRCA1/2 mutation.

In contrast, Sporadic-BC patients exhibited a broader temporal distribution of CVD, with events continuing to accrue beyond the first year of follow-up. This divergence highlights the distinct survivorship trajectories of the two cohorts: BRCA-BC progress rapidly through competing events, while sporadic cases live longer and accumulate cardiovascular morbidity over extended follow-up.

### Fewer observed CVD in BRCA-BC are likely explained by competing risks

Across all analyses, Matched BRCA-BC exhibited an inverse association with the transition from BC diagnosis to the first cardiovascular event. The median pooled hazard ratio (HR) was 0.46 (*p*<0.05 for ∼37% of all subgroups), indicating that Matched BRCA-BC experienced fewer observed CVD compared with MS32 (Figure 3).

**Figure 3:**
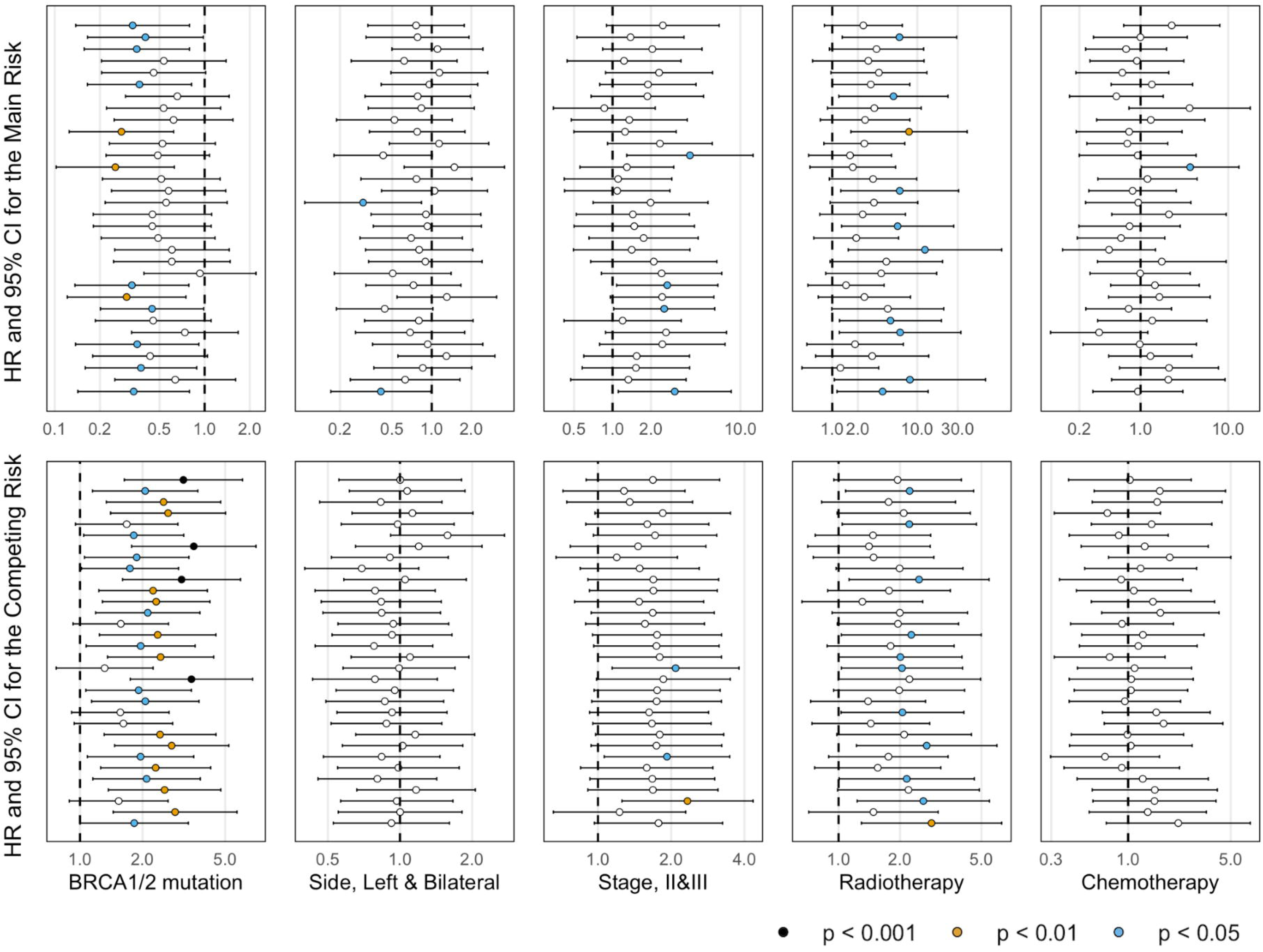
Results of the multi-state Cox proportional hazards regression model. This figure shows the HR (showed with circles) and 95% CI (bar lines), separately for each of the 32 matched subgroups, obtained from the multi-state Cox proportional hazards regression model. The first row presents the results for the transition from BC diagnosis to the first cardiovascular event. The second row shows the results of the competing risks, which represent the transition from BC diagnosis to either the first non-cardiovascular death or distant metastasis. Statistically significant findings are indicated by color-filled circles. To ensure that fine details remain visible, the figures were presented with different x-axis scales.CI=Confidence interval, HR=Hazard ratio.

In contrast, Matched BRCA-BC had a higher hazard of competing events. The transition from BC diagnosis to distant metastasis or non-cardiovascular death was associated with a median HR=2.11 (*p*<0.05 for ∼81% of all subgroups) in Matched BRCA-BC compared with the MS32 (Figure 3). This difference is also evident in the main cohort, as illustrated in Figure 4, which shows that competing events occurred not only more frequently but also at younger ages in the BRCA-BC than in the Sporadic-BC. Because cardiovascular morbidity typically manifests later in life, this early occurrence of competing risks shortened the effective time at risk for CVD, thereby producing the apparent inverse association between BRCA-BC status and cardiovascular outcomes.

**Figure 4.**
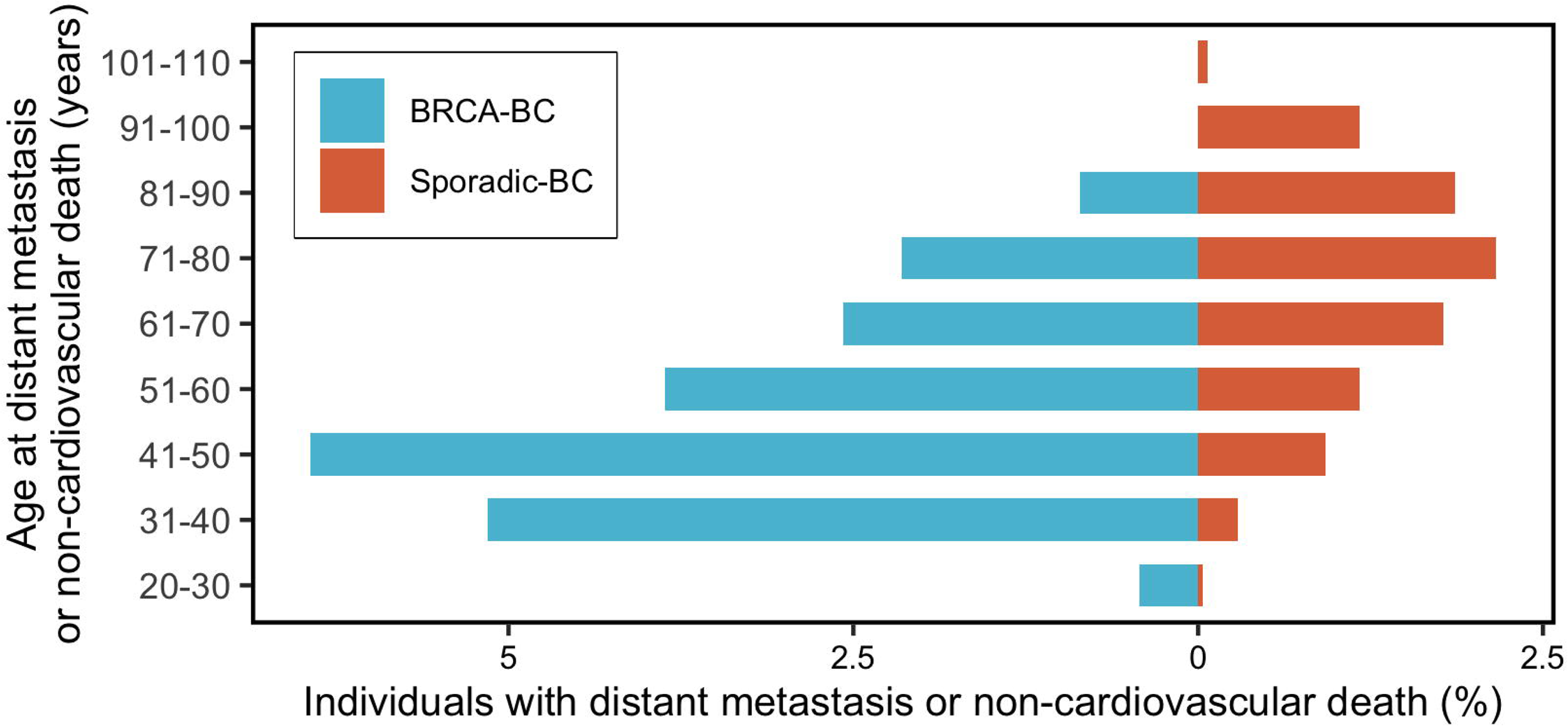
The age distribution of competing risk in the main population. The figure presents the age distribution of individuals in the main population who experienced their first distant metastasis or death. Along with a higher incidence rate among BRCA-BC, a pronounced shift toward younger ages was observed. BRCA-BC=Individuals with BRCA1/2 germline mutations who diagnosed with breast cancer in the main population, Sporadic-BC=Individuals with breast cancer without known status of BRCA1/2 mutation.

The multi-state Cox models also suggested no increased hazards of CVD associated with chemotherapy (median HR=0.98; statistically non-significant for ∼97% of all subgroups). On the other hand, radiotherapy suggested increased hazards of CVD (median HR=3.11; *p*<0.05 for ∼31% of all subgroups). Tumor laterality (left-sided or bilateral versus right-sided disease) was not associated with elevated cardiovascular risk (median HR=0.79; statistically non-significant for ∼94% of all subgroups). Detailed HR and 95% confidence interval (95% CI) for all 32 subgroups are shown in Figure 3.

The additional analyses using logistic regression in the matched subgroups reproduced the inverse association between BRCA-BC status and the risk of experiencing the first CVD (Supplemental Figure 1). Taken together, these results indicate that the lower observed incidence of CVD among BRCA-BC does not imply lower risk for developing CVD. Instead, it most plausibly reflects the earlier occurrence of competing risks, which shorten the period during which CVD can manifest.

## Discussion

We found that BRCA-BC status is associated with a lower observed incidence of first cardiovascular event after BC diagnosis than matched BC survivors without known BRCA1/2 mutations, when accounting for competing risks (distant metastasis, non-cardiovascular death). Simultaneously, Matched BRCA-BC had significantly higher hazards of relapse, metastasis, or death at younger ages, which appears to compress the follow-up interval in which CVD could manifest. Thus, although the hazard ratio for CVD among Matched BRCA-BC is <1, this likely reflects a truncated risk time rather than a true protective biological effect [30].

These findings are consistent with recent studies in the field. For example, Terra et al. (2025) [20], reported no statistically significant increase in risk of ischemic heart disease IHD nor in heart failure among BRCA1/2 mutation carriers compared with non-mutation carriers. Similarly, Demissei et al. (2022) [13] found no significant difference in echocardiographic or cardiopulmonary functional measures between BRCA-BC and non-mutation carriers after doxorubicin treatment of BC. Those studies generally focused on longer-term outcomes, or on functional or subclinical endpoints, and often restricted themselves to survivors with a minimum follow-up period or excluded early post-diagnosis events, which may reduce their sensitivity to early cardiovascular events.

One strength of our study is the absence of a minimum follow-up requirement, which allowed us to observe the early post-diagnosis period, a time characterized by intensive treatment and higher risks of metastasis or death. This allowed observation of the clustering of CVDs in the first year after BC diagnosis (especially among BRCA-BC) better. We also constructed 32 strictly matched 1:1 subgroups to control for pre-existing CVD, CVRFs, tumor stage, age at diagnosis, and laterality, which might help to minimize confounding. Additionally, the use of multi-state Cox models to explicitly recognize competing risks may ensure that estimates for the hazard of CVD properly account for individuals who experience metastasis or death before a CVD could occur. Such modeling could reduce bias compared to considering metastasis or non-cardiovascular death simply as censoring without modeling the competing risk structure. Additionally, this study is population-based and leverages high-quality registry data. The observed first-year clustering of CVD events, particularly among BRCA-BC, highlights a critical window for cardiovascular monitoring, prevention, and intervention that may be underappreciated in studies applying longer minimum follow-up threshold

Overall, current results do not support the hypothesis that BRCA-BC has a higher risk for CVD in the short-to-intermediate period post-BC diagnosis compared with those without known BRCA mutations; however, BRCA-BC highlights important dynamics of competing risk and early cardiovascular event clustering. These findings suggest avenues for future research, particularly to clarify whether specific mutation types, treatment regimens, or genetic modifiers affect cardiovascular risk in BRCA-BC, once sufficient long-term follow-up data becomes available.

### Study limitations

There are limitations that affect interpretation in this study. First, the Sporadic-BC was not systematically tested for BRCA1/2 mutations; some individuals may be undetected mutation carriers, leading to misclassification. That would tend to dilute differences (bias toward the null) rather than produce a false inverse association. Second, although follow-up in some individuals extends to a decade, many have shorter observation periods; because CVD often arises years after treatment, late effects may be under-observed in our sample. Third, our results primarily reflect CVD occurrence during the metastasis-free survivorship window rather than the total post-diagnosis CVD burden, and differential competing events may partly explain the observed inverse association. Fourth, despite matching, residual confounding remains possible, especially for treatment details we could not fully capture (e.g., specific chemotherapy agents, radiotherapy dose fields, internal mammary chain exposure). Fifth, event counts among BRCA-BC were limited, affecting statistical power for detecting more modest hazard ratios or interactions.

### Conclusion

This matched cohort analysis does not support an increased risk for CVD among breast cancer patients who are BRCA1/2 mutation carriers compared with those without known BRCA mutations. On the contrary, we observed an apparent inverse association, which is most plausibly explained by the high incidence of early competing risks, distant metastasis, and non-cardiovascular death, particularly within the first year after diagnosis. This early period, often excluded from prior studies, accounted for a high proportion of cardiovascular events and should be recognized as a critical window in cardio-oncology research.

Beyond this, the findings raise the possibility that a small set of BRCA-BC who survive beyond the early high-risk phase may exhibit distinct biological characteristics, possibly possessing protective mechanisms against CVD. Although our study was not designed to identify such mechanisms, this hypothesis warrants further investigation.

Taken together, these results highlight the importance of analyzing early post-diagnosis cardiovascular outcomes, carefully accounting for competing risks, and considering heterogeneity among BRCA-BC. Future work should focus on long-term follow-up with larger cohorts and detailed genetic characterization to clarify whether distinct subgroups of mutation carriers exist with different cardiovascular risk profiles.

## Author’s contribution

Conceptualization was carried out by L.H., A.L., E.H., and N.A.K. Data acquisition: E.H and A.L. Data curation was performed by N.M., C.L.S., J.R., and E.H. Formal analysis was conducted by M.D.M. Funding acquisition and investigation were undertaken by E.H. and N.A.K. Methodology development involved M.D.M., N.M., A.M., and N.A.K. Project administration and supervision were performed by E.H. and N.A.K. Visualization was performed by M.D.M. M.D.M. and N.A.K. wrote the original draft of the manuscript with critical input from L.H, A.M., and E.H. All authors contributed to review and editing the manuscript.

## Précis

We found that BRCA-associated breast cancer was not associated with an increased risk of cardiovascular disease after diagnosis compared with a matched cohort of sporadic breast cancer. Notably, BRCA1/2-associated patients showed a pronounced spike in cardiovascular events during the first year after diagnosis, together with a high burden of competing mortality.

## Data availability

The data that support the findings of this study are restricted. Access can be requested through the Swedish National Data Service for research purposes, subject to ethical approval.

## Ethical approval

This study was approved by the Swedish Ethical Review Board (2018/2669-31/2, amendment 2019-00540, 2020-07086 and 2024-00960-02) and adhered to the STROBE guidelines and the Declaration of Helsinki. According to Swedish legislation, written informed consent is not required from individuals included in national quality registries for the use of their data in healthcare research.

## Abbreviations list

CVD: Cardiovascular disease
CVRF: Cardiovascular risk factor
BC: Breast cancer
BRCA-BC: Individuals with BRCA1/2 germline mutations who diagnosed with breast cancer in the main population
Sporadic-BC: Individuals with breast cancer without known status of BRCA1/2 mutation
MS32: Collection of 32 matched Sporadic-BC subgroups
HR: Hazard ratio
IQR: Interquartile range
ICD: International Classification of Diseases

